# External Validation of Six Scores Differentiating Atherosclerotic vs. Embolic Large Vessel Occlusion

**DOI:** 10.64898/2026.02.11.26346119

**Authors:** Kenichi Sakuta, Ryoji Nakada, Kenichiro Sakai, Motohiro Okumura, Hiroyuki Kida, Haruhiko Motegi, Gota Nagayama, Rintaro Tachi, Shinji Miyagawa, Teppei Komatsu, Hidetaka Mitsumura, Hiroshi Yaguchi, Yasuyuki Iguchi

## Abstract

**Purpose:** Intracranial atherosclerotic disease-related large vessel occlusion (ICAD-LVO) presents distinct challenges, particularly regarding the high risk of reocclusion and the need for specific management strategies. While several prediction scores exist to differentiate ICAD-LVO from embolic LVO (EMB-LVO), their external validity remains unproven. We aimed to externally validate six established prediction scores for differentiating the two.

**Methods:** We analyzed data from a prospectively maintained, two-center stroke registry (June 2021–March 2025). Consecutive patients who underwent mechanical thrombectomy and had complete clinical and imaging data necessary for calculating six scores (ISAT, REMIT, ABC^2^D, ATHE, ICAS-LVO, and Score-ICAD) were included. LVO etiology was defined based on angiographic findings during endovascular treatment. The discriminative performance of each score was assessed using the area under the receiver operating characteristic curve (AUC).

**Results:** Of 1,288 screened admissions, 91 patients met the inclusion criteria (ICAD-LVO, n = 18; embolic occlusion, n = 73). The AUCs (95% confidence interval) for differentiating etiology were: ISAT, 0.870 (0.664–1.000; P = 0.064); REMIT, 0.793 (0.676–0.911; P <0.001); Score-ICAD, 0.707 (0.582–0.833; P = 0.013); ABC^2^D, 0.627 (0.504–0.751; P = 0.095); ATHE, 0.600 (0.451–0.749; P = 0.230); and ICAS-LVO, 0.465 (0.301–0.630; P = 0.650).

**Conclusion:** In this external validation, REMIT demonstrated the most robust and statistically significant discrimination between ICAD-LVO and EMB-LVO. Overall, scores incorporating imaging features outperformed those relying on clinical variables. These findings support the concept that ICAD-LVO represents a distinct pathophysiological entity from embolic occlusion and that accurate mechanism inference requires comprehensive imaging assessment of intracranial atherosclerotic disease beyond the occlusion site.

## Introduction

Mechanical thrombectomy has become the established standard of care for treating acute ischemic stroke caused by large vessel occlusion (LVO) ^1^. However, LVO etiologies are heterogeneous, presenting distinct challenges depending on whether the occlusion is embolic or related to intracranial atherosclerotic disease (ICAD) ^2^. Residual stenosis and reocclusion may occur after treatment of ICAD-related LVO (ICAD-LVO), necessitating rescue therapies such as angioplasty, stenting, or antiplatelet administration^3^. Consequently, accurate identification of the occlusion mechanism before or during the procedure is pivotal for optimizing treatment strategies ^4,5^. Despite this clinical need, differentiating ICAD-LVO from embolic LVO (EMB-LVO) in the acute setting remains a diagnostic challenge.

To address this challenge, several prediction scales have been proposed to differentiate the two by integrating clinical variables and imaging findings ^2^. These scales encompass diverse factors, ranging from demographic characteristics and atrial fibrillation to vascular morphology on computed tomography angiography (CTA) and infarct patterns ^6,7^. However, substantial heterogeneity exists among these prediction instruments regarding target vascular territories (anterior vs. posterior), specific variables selected, and the weighting of clinical versus imaging components^8–10^. Furthermore, as most scales were developed in restricted derivation cohorts, their generalizability has not yet been fully established. Crucially, the relative contribution of individual scale components to prediction of LVO etiology has not been systematically scrutinized.

The primary objective of this study was to externally validate existing prediction scales for differentiating ICAD-LVO from EMB-LVO using a large multicenter consecutive stroke registry. Furthermore, by dissecting each scale into its individual components, we sought to determine the independent predictive value of each factor and isolate the key elements driving discrimination of LVO etiologies in the acute setting. Through this comprehensive analysis, we aimed to not only verify the generalizability of current scales, but also to provide critical insights to develop more practical and pathophysiologically grounded prediction models.

## Methods

### Study Design and Setting

This was a retrospective analysis of prospectively collected data from a multicenter stroke registry. The registry includes consecutive patients with acute stroke admitted to Jikei University Hospital and Jikei University Kashiwa Hospital. The study period was from June 2021 to March 2025, which was selected to avoid overlap with the cohort used for development of the REMIT scale^7^. Eligible patients were consecutive adults with acute ischemic stroke due to LVO who underwent mechanical thrombectomy. Inclusion criteria were: (1) acute ischemic stroke caused by LVO, (2) treatment with mechanical thrombectomy, and (3) availability of complete clinical and imaging data required for calculation of all study scores. Exclusion criteria were: (1) inability to undergo contrast-enhanced computed tomography, (2) presence of tandem lesions, and (3) availability of only preprocedural magnetic resonance imaging. Patients who could not or did not undergo contrast-enhanced computed tomography were excluded because such data were required for prediction scale scoring. Those with tandem lesions were excluded because such complex pathology precludes standardized scoring and may confound classification. ICAD-LVO was defined as >50% residual stenosis at the culprit site after thrombectomy, as in previous studies^8,11^. Patients who did not meet the criteria for ICAD-LVO were classified as EMB-LVO.

A literature search was conducted in PubMed using the following search terms: (Large Vessel Occlusion OR LVO) AND (Intracranial Atherosclerosis OR ICAS OR ICAD) AND (Prediction OR Score OR Scale OR Nomogram) AND (Embolism OR Embolic) AND (Differentiate OR Predict). The search identified six studies that proposed scales for predicting the etiology of large vessel occlusion^6–11^ (Table 1). For calculation of the six scales, the following imaging features were assessed: hyperdense arterial sign ^12^, truncal-type occlusion^13^, tapered sign^14^, non-culprit stenosis^15^, border zone infarction, cortical infarction, calcification at the carotid siphon, and multiple arterial stenoses^6^. The operational definitions of each imaging sign were as follows: The hyperdense arterial sign was defined as intraluminal arterial hyperattenuation compared with the contralateral corresponding artery and the adjacent brain parenchyma. Truncal-type occlusion was defined on CTA as one in which the major distal bifurcation (e.g., the middle cerebral artery bifurcation) was clearly visualized, indicating that the occlusion was located in the proximal arterial trunk rather than at the bifurcation site. The tapered sign was defined as an occlusion morphology characterized by gradual narrowing of contrast opacification toward the occlusion site, ending in a tapered, sharp configuration. Non-culprit stenosis was defined as a 50% to 99% stenosis identified in intracranial arteries other than the occluded artery. Border zone infarction was defined as superficial cortical or deep internal border zone territory infarction. Cortical infarction was defined as any acute, subacute, or chronic cortical infarction located distal to the region of stenosis within the territory supplied by the corresponding intracranial artery, excluding cortical border zone areas. Calcification at the carotid siphon was defined as calcification detected on admission CTA at the ipsilateral internal carotid artery siphon, specifically involving the supraclinoid segment or the communicating portion immediately proximal to the carotid bifurcation. Multiple arterial stenosis was defined as the presence of ≥50% stenosis in two or more intracranial arteries other than the occluded vessel.

**Table 1.**
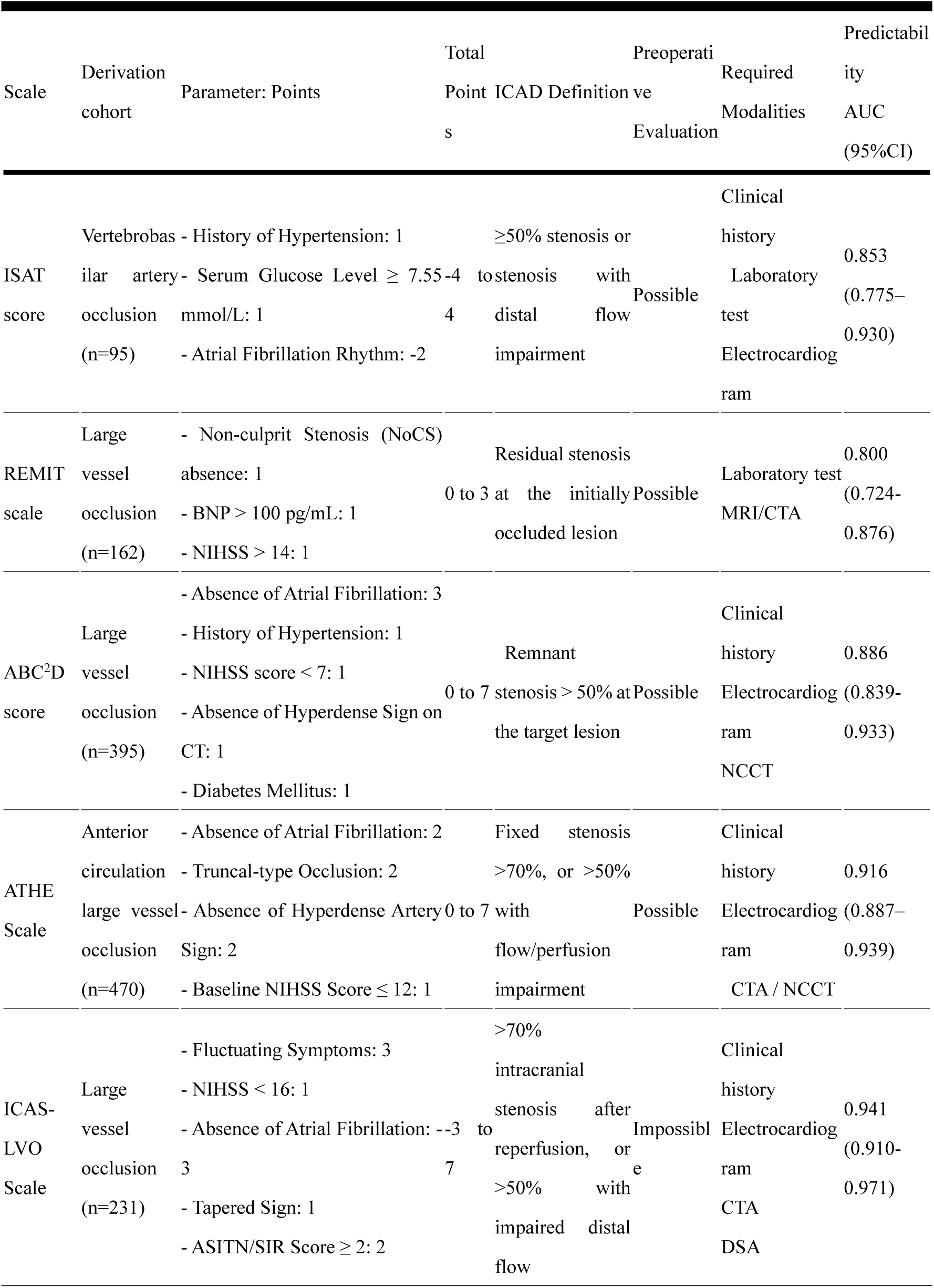
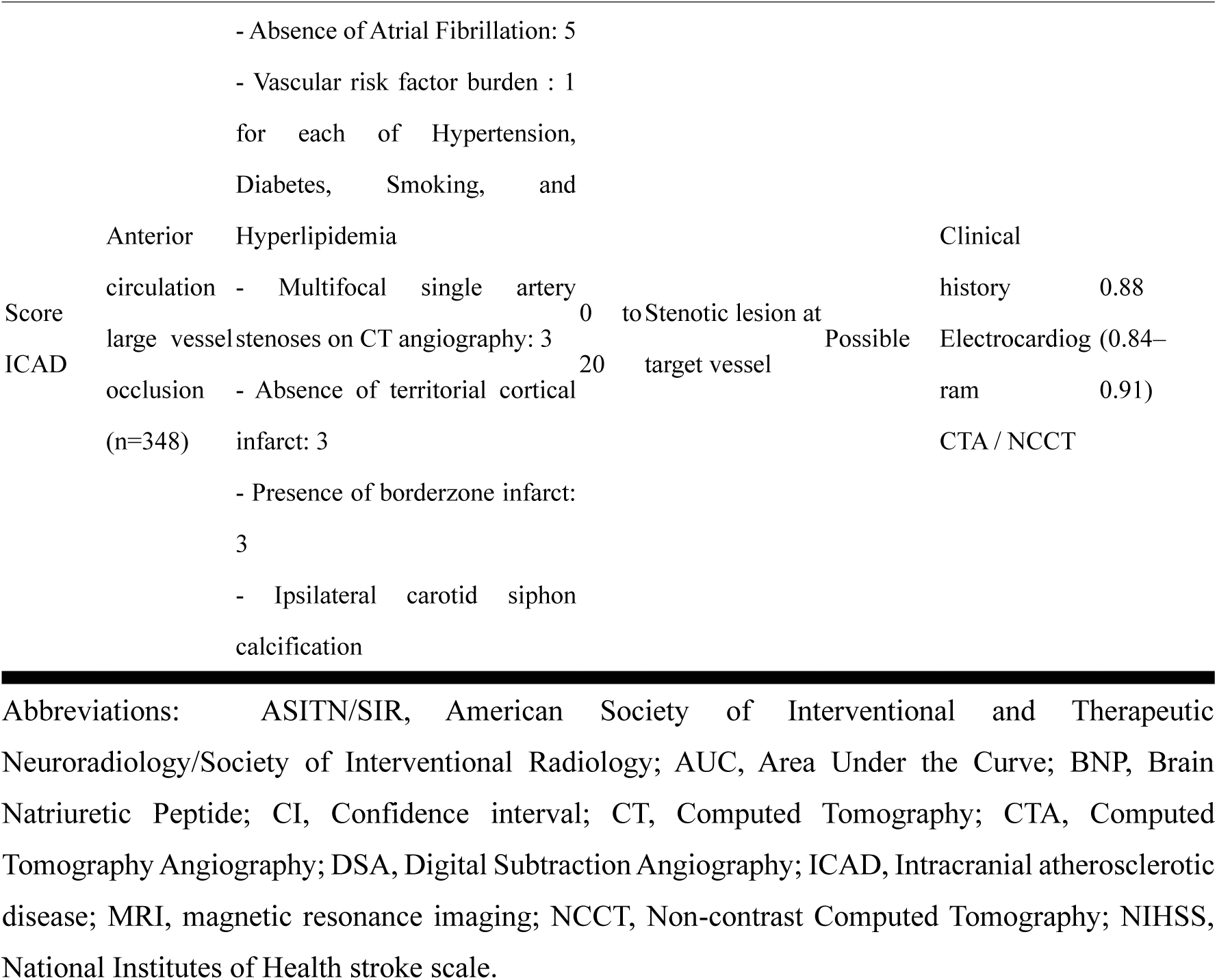
Comparative Characteristics of Six Etiology Prediction Scales for Atherosclerotic vs. Embolic Large Vessel Occlusion Prior to Mechanical Thrombectomy.

This study was approved by the Ethics Committee of Jikei University School of Medicine (approval numbers: 31–169 [9668] and 29–196 [8812]). Informed consent was obtained using an opt-out approach. All study procedures were conducted in accordance with the Declaration of Helsinki.

### Clinical Background

The following clinical variables were collected: age, sex, body mass index, length of hospital stay, and laboratory data obtained on admission, including estimated glomerular filtration rate, glycated hemoglobin, and concentrations of serum glucose, brain natriuretic peptide (BNP), low-density lipoprotein cholesterol, and D-dimer. Traditional cardiovascular risk factors were also recorded, including diabetes mellitus, hypertension, dyslipidemia, atrial fibrillation, ischemic heart disease, peripheral arterial disease, malignant neoplasms, and chronic kidney disease. Stroke severity was assessed on admission by a neurologist using the National Institutes of Health Stroke Scale (NIHSS).

### Statistical Analyses

Baseline characteristics of the study cohort were summarized using medians with interquartile range for continuous variables and counts with percentage for categorical variables. Patients were grouped according to LVO etiology. Intergroup comparisons were performed using the chi-square test or Fisher’s exact test for categorical variables and the Student’s t test or Mann–Whitney U test for continuous variables, as appropriate. All tests were two-sided. P <0.05 was considered significant.

Each prediction scale was then calculated, and its discriminatory performance was assessed using the area under the receiver operating characteristic curve (AUC). Complete case analysis was performed. In accordance with the design characteristics of each scale, AUCs were calculated for the following target populations: the REMIT scale for EMB-LVO, the ISAT score in patients with posterior circulation occlusion only, and the ATHE score and Score-ICAD in patients with anterior circulation occlusion only. Finally, all individual components included in the six prediction scales were extracted, and their associations with ICAD-LVO were evaluated by calculating odds ratios with 95% confidence intervals (CIs). Statistical analyses were performed using SPSS software version 22 (IBM Corp., Armonk, NY, USA).

## Results

Among 1,288 consecutive patients with acute ischemic stroke, 91 patients met inclusion criteria (Figure 1). Patient characteristics are summarized in Table 2. ICAD-LVO was identified in 18 patients (20%), and 87% of LVOs involved the anterior circulation. The prevalence of atrial fibrillation (11% vs. 41%) and BNP concentration (55.1 pg/mL vs. 121.4 pg/mL) were significantly lower in the ICAD-LVO group than the EMB-LVO group. Among the 18 patients with posterior circulation LVO, three were diagnosed with ICAD-LVO.

**Figure 1.**
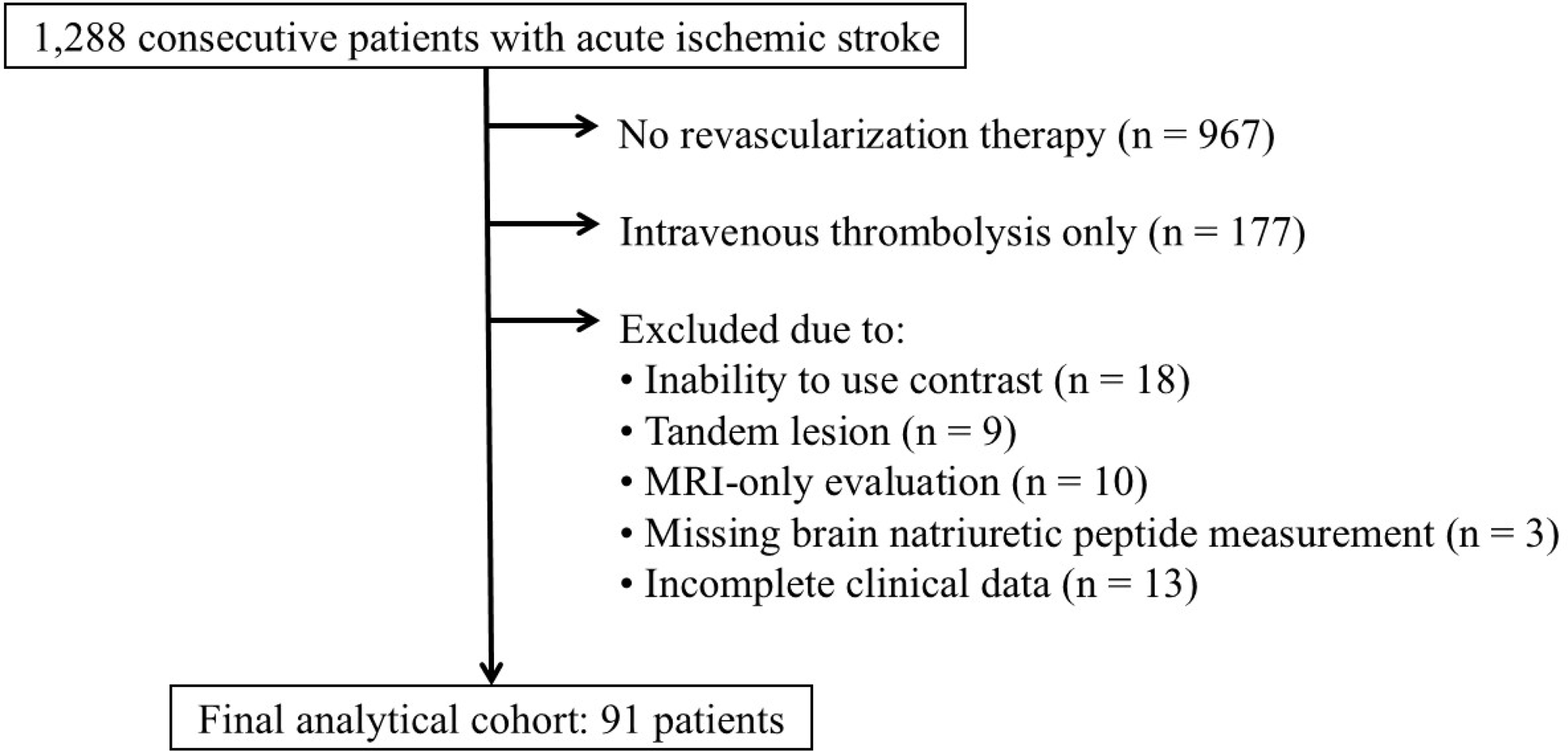
Flow diagram of patient selection

**Table 2.** Patient characteristics.

|  | total<br>(n=91) | ICAD-LVO<br>(n=18) | EMB-LVO<br>(n=73) | P Value |
| --- | --- | --- | --- | --- |
| Demographics |  |  |  |  |
| Men, n(%) | 52 (57) | 11 (61) | 41 (56) | 0.704 |
| age, median(IQR) | 78 (68-82) | 73 (68-80) | 79 (68-82) | 0.304 |
| Medical history, n(%) |  |  |  |  |
| Diabetes Mellitus | 18 (20) | 1 (6) | 17 (23) | 0.079 |
| Hypertension | 49 (54) | 11 (61) | 38 (52) | 0.490 |
| Dyslipidemia | 23 (25) | 3 (17) | 20 (27) | 0.270 |
| Atrial fibrillation | 32 (35) | 2 (11) | 30 (41) | 0.017 |
| Ischemic heart disease | 22 (24) | 2 (11) | 20 (27) | 0.125 |
| Stroke | 13 (14) | 2 (11) | 11 (15) | 0.502 |
| Dementia | 9 (10) | 1 (6) | 8 (11) | 0.433 |
| Chronic kidney disease | 6 (7) | 0 (0) | 6 (8) | 0.255 |
| Malignant neoplasms | 23 (25) | 4 (22) | 19 (26) | 0.501 |
| Physical exam, median(IQR) |  |  |  |  |
| BMI | 21.9 (19.9-24.2) | 22.0 (20.5-24.0) | 21.9 (19.8-24.3) | 0.955 |
| Systolic blood pressure, mmHg | 151 (131-178) | 168 (153-178) | 146 (130-177) | 0.168 |
| Diastolic blood pressure, mmHg | 86 (70-100) | 91 (76-100) | 84 (70-99) | 0.532 |
| Heart rate, /minute | 84 (72-96) | 81 (71-90) | 84 (73-98) | 0.276 |
| NIHSS | 19 (12-26) | 19 (13-26) | 18 (11-25) | 0.881 |
| Laboratories, median(IQR) |  |  |  |  |
| Glucose, mg/dL | 123 (113-161) | 123 (108-163) | 130 (114-156) | 0.687 |
| HbA1c, % | 5.8 (5.6-6.2) | 5.7 (5.4-5.9) | 5.9 (5.6-6.3) | 0.102 |
| eGFR, mL/min/1.73 m<sup>2</sup> | 59.6 (45.9-77.4) | 71.1 (48.2-85.4) | 57.6 (45.0-75.7) | 0.200 |
| BNP, pg/mL | 107.9 (49.0-353) | 55.1 (18.5-219.3) | 121.4 (52.6-385.8) | 0.037 |
| LDL-C, mg/dL | 105 (86-124) | 114 (94-132) | 103 (85-122) | 0.172 |
| D-dimer, µg/mL | 1.8 (1.1-4.1) | 2.1 (1.0-3.7) | 1.8 (1.1-4.3) | 0.906 |
| Anterior circulation, % | 79 (87) | 15 (83) | 64 (88) | 0.437 |
| Intravenous thrombolysis, % | 53 (58) | 9 (50) | 44 (60) | 0.429 |
| Hospital stay, day, median(IQR) | 27 (10-45) | 33 (22-56) | 25 (10-44) | 0.137 |

Abbreviations: BMI, Body Mass Index; BNP, Brain Natriuretic Peptide; eGFR, estimated Glomerular Filtration Rate; EMB-LVO, embolic large vessel occlusion; ICAD-LVO, Intracranial atherosclerotic disease-related large vessel occlusion; IQR, interquartile range; LDL-C, Low Density Lipoprotein Cholesterol; NIHSS, National Institutes of Health stroke scale.

Figure 2 displays the receiver operating characteristic analyses for predicting ICAD-LVO. The ISAT score yielded the highest AUC of 0.870 (95% CI: 0.664–1.000), showing a trend toward significance (P = 0.064). The REMIT scale followed with a robust and statistically significant AUC of 0.793 (95% CI: 0.676–0.911, P <0.001). The Score-ICAD also proved to be a significant predictor, yielding an AUC of 0.707 (95% CI: 0.582–0.833, P = 0.013). Conversely, the other three scales (ABC^2^D, ATHE, and ICAS-LVO) showed poor discrimination for ICAD-LVO in our dataset. Specifically, the ABC^2^D and ATHE scales had AUCs of 0.627 (95% CI: 0.504-751, P = 0.095) and 0.600 (95% CI: 0.451-0.749, P = 0.230), respectively, while the ICAS-LVO scale showed an AUC of 0.465 (95% CI: 0.301-0.630, P = 0.650), indicating no predictive value.

**Figure 2.**
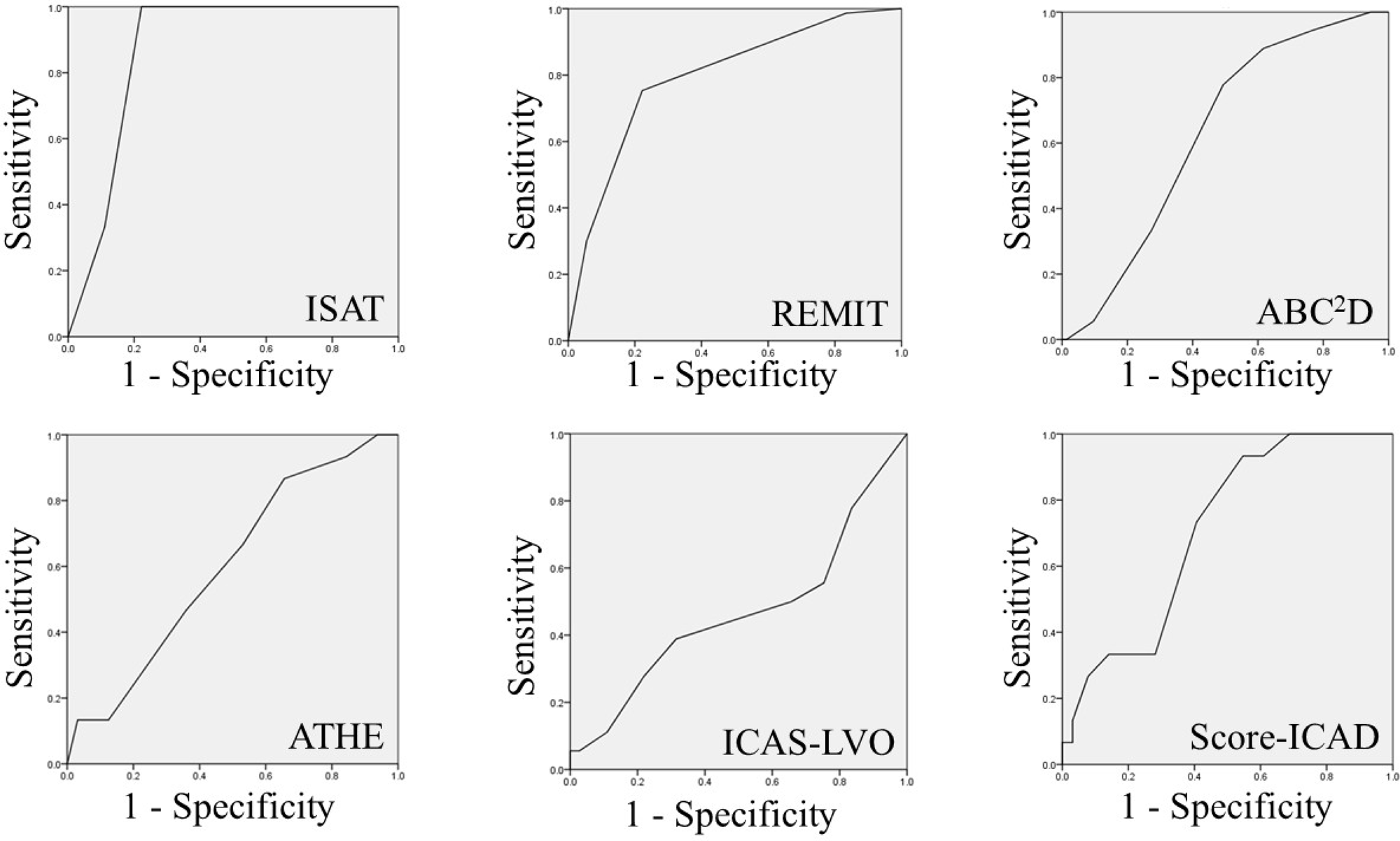
Receiver operating characteristic curves for prediction of intracranial atherosclerotic disease–related large vessel occlusion (ICAD-LVO). AUCs (95% CI) were: ISAT, 0.870 (0.664–1.000); REMIT, 0.793 (0.676–0.911); ABC^2^D, 0.627 (0.504–0.751); ATHE, 0.600 (0.451–0.749); ICAS-LVO, 0.465 (0.301–0.630); Score-ICAD, 0.707 (0.582–0.833).

The results of the analysis evaluating the predictive ability of individual components included in the six prediction scales for ICAD-LVO are summarized in Table 3. Among variables related to medical history, the absence of atrial fibrillation was the only factor significantly associated with ICAD-LVO. Among physical examination findings and laboratory variables, no factors were significantly predictive of ICAD-LVO. In contrast, among imaging findings, the tapered sign, non-culprit stenosis, border zone infarction, and multiple arterial stenosis were significantly associated with ICAD-LVO. Of all the variables examined, non-culprit stenosis demonstrated the highest odds ratio for predicting ICAD-LVO.

**Table 3.**
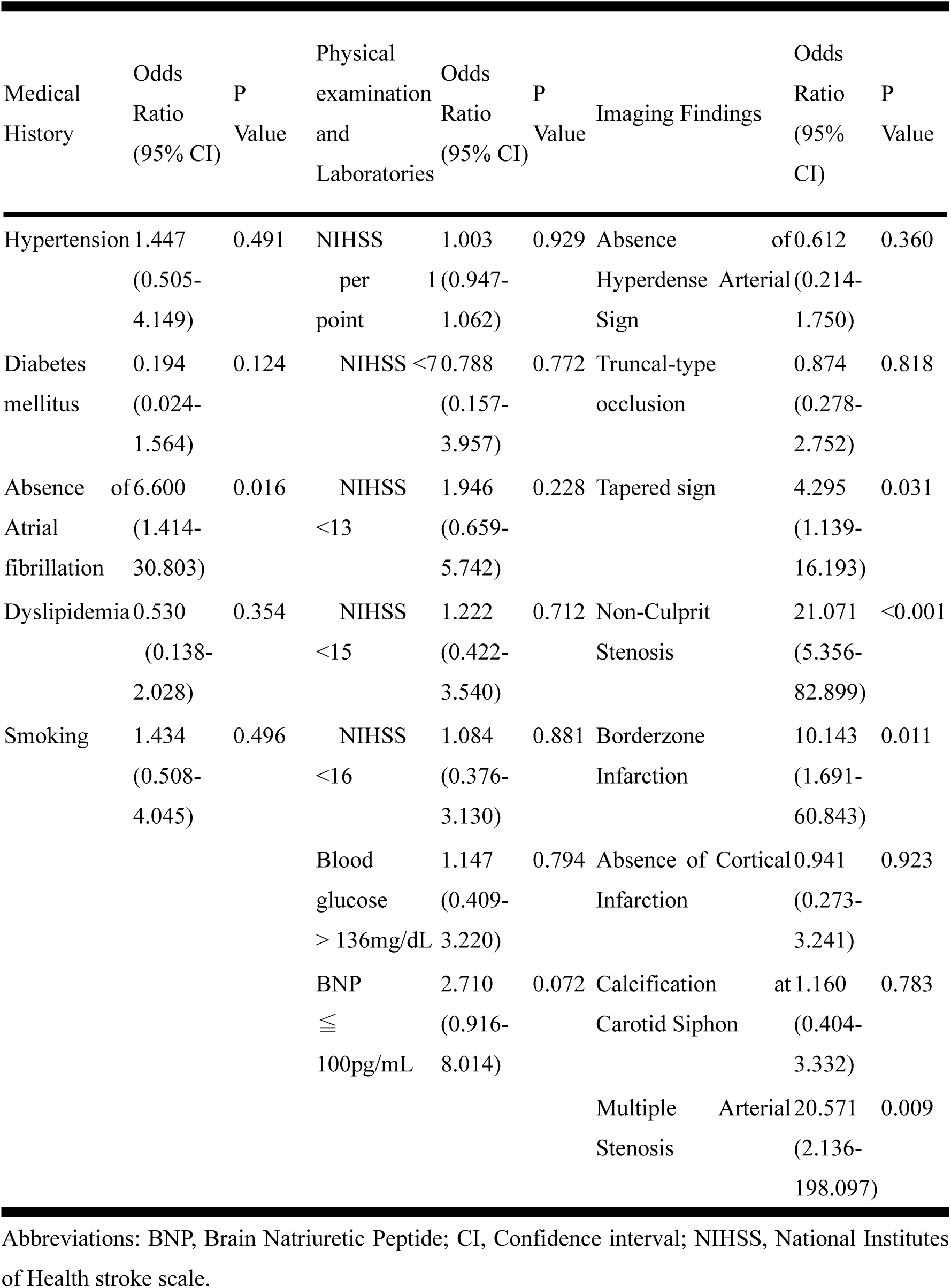
Univariate Odds Ratios of Factors Derived from Six Etiology Prediction Scales for ICAS vs. Embolic LVO Prior to Thrombectomy.

## Discussion

In this multicenter study using a consecutive stroke registry, we performed an external validation of six previously proposed prediction scales designed to differentiate ICAD-LVO from EMB-LVO. In addition to evaluating the overall discriminatory performance of each scale, we further dissected their individual components to clarify which factors most strongly contributed to etiology prediction in an independent cohort. Notably, imaging findings were more strongly associated with ICAD-LVO than non-imaging clinical variables, such as atrial fibrillation and NIHSS score, highlighting the central role of angiographic and neuroimaging features in distinguishing ICAD-LVO in the acute setting.

Only two prediction scales, the REMIT scale and Score-ICAD, demonstrated statistically significant predictive performance for ICAD-LVO. The performance of the prediction scales observed in this study differed from that reported in their original derivation studies. Each scale has distinct characteristics, as outlined below. The ISAT score was developed exclusively for posterior circulation occlusion and does not incorporate imaging findings^8^. The REMIT scale places greater emphasis on clinical variables associated with embolic mechanisms and has the advantage of not requiring differentiation between computed tomography- and magnetic resonance-based imaging findings^7^. The ABC^2^D score is largely driven by clinical data ^11^. The ATHE score was designed for anterior circulation occlusion and assigns substantial weight to imaging features^9^. The ICAS-LVO scale is characterized by the requirement for intraprocedural angiographic findings for accurate score calculation^10^. The Score-ICAD places strong emphasis on imaging findings that reflect intracranial atherosclerotic disease^6^. These differences in target populations and in the weighting of clinical and imaging variables may have contributed to the variability in discriminatory performance observed in the present cohort. Because the analyzed cohorts differed across scales, direct comparisons of AUCs should be interpreted with caution. In particular, the results for posterior circulation occlusion, in which the number of cases was limited, warrant careful interpretation. Furthermore, the attenuated performance observed in this study may reflect the inherently conservative nature of external validation, in which predictive accuracy is typically lower than that reported in derivation cohorts^16^.

Imaging findings demonstrated stronger predictive value for ICAD-LVO than non-imaging clinical variables such as atrial fibrillation, NIHSS score, and BNP concentration. This finding suggests that ICAD-LVO is primarily characterized by pathological features intrinsic to the occlusion site itself rather than by systemic clinical background factors. ICAD-LVO typically arises from in situ thrombosis on pre-existing atherosclerotic stenosis and is often accompanied by chronic hypoperfusion, resulting in characteristic vascular morphology and infarct distribution that can be directly captured on neuroimaging^17–20^. In contrast, clinical variables such as atrial fibrillation and BNP concentration, while informative for identifying embolic etiology, may provide only indirect information when attempting to identify ICAD-LVO. Notably, imaging predictors that were significantly associated with ICAD-LVO in this study—including non-culprit stenosis, tapered sign, border zone infarction, and multiple arterial stenosis—collectively reflect a diffuse atherosclerotic burden and chronic hemodynamic compromise beyond the culprit lesion alone^15,19,21^. These findings support the concept that ICAD-LVO represents a disease process occurring along a continuum of intracranial atherosclerosis rather than an isolated, abrupt occlusive event. Recent studies have further suggested that perfusion imaging may provide complementary mechanistic insights into ICAD-LVO^2^. Perfusion-based parameters such as mismatch ratios, Tmax profiles, and preserved or increased cerebral blood volume reportedly reflect adaptive collateral circulation and vascular reserve associated with chronic intracranial stenosis ^22–24^. Accordingly, future prediction models that integrate conventional morphological imaging findings with perfusion-based metrics may enable more accurate and pathophysiologically grounded identification of ICAD-LVO in the acute setting.

This study has several limitations. First, selection bias and unmeasured confounding may have been present, as in any retrospective registry study. Second, the number of ICAD-LVO cases was modest, particularly in the posterior circulation, which may have limited the statistical power of certain analyses. Consequently, although ISAT demonstrated a high AUC, the association did not reach statistical significance. Third, defining ICAD-LVO based on angiographic findings rather than pathological confirmation may have introduced misclassification bias. Fourth, as the cohort was derived from a small number of institutions from a single country, our findings may not be generalizable to other populations. Fifth, given our focus on externally validating established scales, we did not attempt to derive a new scoring system or recalibrate variable weights. Finally, the inconsistent availability of perfusion imaging precluded a comprehensive assessment of perfusion-based markers across the entire cohort.

## Conclusion

Current prediction scales have limited accuracy in identifying ICAD-LVO; however, imaging predictors appear to be superior to clinical factors. ICAD-LVO represents a distinct pathophysiological entity from embolic occlusion, and accurate mechanism inference requires comprehensive imaging assessment of the extent of intracranial atherosclerotic disease, encompassing both the occluded segment and other intracranial arterial territories.

## Data Availability

The data that support the findings of this study are available from the corresponding author upon reasonable request.

## Acknowledgments

None

## Contributors

KSaku performed endovascular procedures, framed the study concept, collected data, and wrote and revised this manuscript. RN and HY offered scientific advice for the concept, data collection, and interpreting of data. MO, HK, MH, GN, RT, SM, TK, KSaka and HM collected data. YI offered scientific advice for the concept, interpretation of data, and revision of the manuscript.

